# Testing Bidirectional Genetic Links Between ADHD and the Big Five Personality Traits

**DOI:** 10.64898/2026.10.05.26363336

**Authors:** Hugo Peyre, Nicolas Hoertel, Baptiste Pignon, Sebastien Weibel, Mario Speranza, Boris Chaumette, Ted Schwaba, Camille M. Williams

**Affiliations:** Autism Reference Centre of Languedoc-Roussillon CRA-LR, Excellence Centre for Autism and Neurodevelopmental disorders CeAND, Montpellier University Hospital, MUSE University, Montpellier, France; CESP Centre de recherche en Épidémiologie et Santé des Populations, INSERM U1178, Villejuif, France; Laboratoire de Sciences Cognitives et Psycholinguistique (ENS, EHESS, CNRS), Ecole Normale Supérieure, PSL University, Paris, France; Université Paris Cité, Inserm U1266, Institut de Psychiatrie et Neuroscience de Paris, Paris, France; Service de Psychiatrie et Addictologie, DMU Psychiatrie et Addictologie, AP-HP, Hôpital Corentin-Celton, Issy-les-Moulineaux, 92130, France; Département Médico Universitaire (DMU) - Innovation en santé Mentale, Psychiatrie et AddiCTologie, Hôpitaux Universitaires « H. Mondor », Assistance Publique hôpitaux de Paris (AP-HP), Université Paris-Est-Créteil (UPEC), Mondor Biomedical Research Institute (IMRB - Inserm), Translational Neuropsychiatry, Fondation FondaMental, Créteil, France; Psychiatry, Mental Health and Addictology Department, University Hospitals of Strasbourg, 1 place de l’Hôpital, Strasbourg, 67000, France; INSERM UMR-S 1329, Strasbourg Translational Neuroscience & Psychiatry, Strasbourg, France; Faculty of Medicine, Strasbourg University, Strasbourg, 67000, France; Versailles Hospital Center, University Department of Child and Adolescent Psychiatry, Le Chesnay 78150, France; Department of Psychiatry, McGill University, Montreal, Canada; Université Paris Cité, NeuroDiderot (INSERM U1141), Institut Pasteur (Human Genetics and Cognitive Functions, CNRS UMR3571), Paris, France; GHU-Paris Psychiatrie et Neurosciences, Hôpital Sainte Anne, Paris F-75014, France; Department of Psychology, Michigan State University, East Lansing, United States

## Abstract

Attention Deficit Hyperactivity Disorder (ADHD) frequently co-occurs with distinct personality profiles, but whether ADHD shapes personality (scar model) or personality influences the development of ADHD (vulnerability model) remains unclear. Leveraging genome-wide association study (GWAS) data, we examined directional associations between ADHD and the Big Five personality traits (Neuroticism, Extraversion, Openness, Agreeableness, and Conscientiousness). We estimated genetic correlations and conducted bidirectional two-sample Mendelian Randomization (MR) analyses between ADHD and the Big Five. Robustness was evaluated using multiple sensitivity tests, including MR-CAUSE, which accounts for correlated and uncorrelated pleiotropy, and colocalization analyses on fine-mapped variants. Higher genetic liability to Neuroticism and Extraversion, alongside lower liability to Conscientiousness, increased ADHD risk, consistent with the vulnerability model. Conversely, ADHD genetic liability was associated with lower Agreeableness, supporting the scar model. Associations were generally robust across sensitivity analyses, with MR-CAUSE supporting the Neuroticism and Agreeableness associations and colocalization providing variant-level support for associations between ADHD and Neuroticism, Conscientiousness, and Agreeableness. These findings provide early, population-level insight into personality-related vulnerability to ADHD and motivate further work on the role of personality traits in ADHD risk.

## INTRODUCTION

Does having Attention Deficit/Hyperactivity Disorder (ADHD) influence a child’s personality, or does their personality make them more susceptible to developing ADHD? These questions are frequently raised by parents in clinical settings and have long been of interest to researchers and clinicians [1–3]. Decades of research support robust associations between ADHD and various personality traits, which are thought to arise from a multitude of genetic and environmental factors [4]. Yet the directionality of these relationships remains unclear.

ADHD is a neurodevelopmental disorder characterized by persistent inattention, hyperactivity, and impulsivity that interferes with daily functioning and development. It is one of the most commonly diagnosed psychiatric disorders in childhood, affecting about 5% of individuals, and persisting into adulthood in approximately 60% of cases [5, 6]. While ADHD is clinically defined as a disorder, its core symptoms are dimensionally distributed in the population [7]. Accordingly, ADHD shows systematic associations with broader personality traits [2], which represent stable, nonclinical dimensions of individual differences.

The Big Five personality traits describe relatively stable individual differences in affect, behavior, and cognition along five broad and mostly independent dimensions: Neuroticism (tendency to experience psychological stress), Agreeableness (trust and altruism), Extraversion (sociability and cheerfulness), Conscientiousness (goal-directed behavior and adherence to principles), and Openness to Experience (intellectual curiosity and willingness to explore new ideas) [8]. In samples of children, personality is typically conceptualized in terms of temperament [9]. However, research suggests a very high degree of overlap between dimensions of temperament and the Big Five, such that childhood personality can also be described in terms of the Big Five [10–12].

Meta-analytic evidence supports strong phenotypic associations between ADHD and certain Big Five dimensions [4, 13, 14]: children and adults with ADHD tend to exhibit higher levels of Neuroticism and lower levels of Conscientiousness and Agreeableness, while Extraversion and Openness show small to no association.

Two frameworks might explain the link between ADHD and personality traits: the scar model and the vulnerability model. The scar model compares the lasting impact of ADHD-related difficulties to a scar that remains on an individual’s personality [15, 16]. This model is consistent with longitudinal studies showing that children with ADHD tend to have higher Neuroticism scores and lower Agreeableness and Conscientiousness scores in adolescence [17]. Conversely, the vulnerability model proposes that certain personality traits, such as high Neuroticism or low Conscientiousness, may increase the risk of developing ADHD [1, 17]. However, it remains unclear whether these associations reflect causal effects of ADHD on personality, vulnerability to ADHD driven by personality, or bidirectional processes.

The phenotypic overlap between ADHD and certain personality traits may stem from environmental factors, such as early childhood experiences, family dynamics, and social interactions [18, 19], as well as neurobiological mechanisms [1, 20]. Both ADHD and personality traits are heritable, indicating that genetic differences contribute to individual variation in these traits [21, 22]. Twin and population studies report moderate genetic overlap between ADHD and personality traits, particularly for Neuroticism (*r*g = 0.24 - 0.56 [22, 23]). While twin studies provide valuable insights into shared genetic influences, they do not provide information on the direction of these relationships, nor do they pinpoint overlapping genetic variants between traits.

Genome-wide association studies (GWAS) allow for a more precise examination of these relationships at the level of common genetic variants, offering new insights into the genetic architecture of ADHD and personality traits. Two-sample Mendelian Randomization (MR) leverages GWAS data to infer causal relationships between traits, establishing whether one trait influences another [24, 25]. While MR has been used to explore causal links between ADHD and various mental health disorders [26], as well as between personality measures and anxiety and depression [22], bidirectional associations between ADHD and the Big Five have only been investigated for Neuroticism and Extraversion. One study reported an effect of Neuroticism on ADHD [27], while another study found that Extraversion exerts an effect on ADHD [28]. However, these studies, particularly the one on Extraversion, were based on GWAS that lacked the statistical power to reliably test these associations. In light of a more recent, larger-scale GWAS release on personality traits [14], we can now investigate the bidirectional association between ADHD and all Big Five personality traits using better powered GWAS and complementary approaches, such as colocalization.

Colocalization complements MR by testing whether associations with the exposure and outcome within a genomic region are likely to reflect the same underlying causal signal [29]. An MR association could otherwise arise from two separate variants correlated through linkage disequilibrium (LD) rather than from a single shared causal variant. Colocalization analyses can now be performed on fine-mapped common genetic variants identified with Sum of Single Effects (SuSiE), which detects credible sets representing distinct association signals [30]. SuSiE-based colocalization then estimates the probability that signals for two traits share an underlying causal variant. Although colocalization may not identify the exact causal variant, it can show that the two association signals are more consistent with sharing an underlying causal variant than with arising from separate variants in LD. This increases confidence that the MR finding is not an LD artifact. Building on these methodological advances, we leverage the largest and latest available GWAS data to investigate the bidirectional relationship between ADHD and each of the Big Five personality traits. After estimating genetic correlations between our traits, we conduct two-sample MR to test for bidirectional relationships between ADHD and each personality dimension. We perform a series of sensitivity tests to evaluate the robustness of the MR results, including colocalization analyses. By integrating multiple approaches, our study addresses a long-standing clinically relevant question: Does ADHD liability influence the development of one’s personality, or do certain personality traits increase one’s risk of developing ADHD?

## METHODS

All statistical analyses were conducted using R version 4.0.3 and 4.5.1 [31].

### GWAS selection

We included the largest and most recent public summary statistics. The Big Five GWAS were provided by the Revived Genomics of Personality Consortium (ReGPC) on approximately 500 000 to 1 million participants from European-like ancestry groups [14] and the ADHD GWAS conducted on 38 691 cases and 186 843 controls from European-like ancestry groups [21] was downloaded from the Psychiatric Genomics Consortium. ADHD cases were diagnosed in childhood or adolescence based on ICD-10 criteria [21].

### Genetic correlation analyses

We estimated genetic correlations between the munged ADHD and the Big Five personality traits using the *munge* and *ldsc* (linkage disequilibrium score regression) functions in the GenomicSEM R v0.0.5 package [32]. GWAS were munged using HapMap3 SNPs from European samples with an info filter of ≥ 0.9 and a minor allele frequency (MAF) threshold of ≥ 0.01. We ran LDSC using LD scores calculated from the 1000 Genomes Project Phase 3 European superpopulation [33], filtered for MAF ≥ 0.005. Population and sample prevalence were set to NA (not available) for the continuous personality traits. For the ADHD GWAS, the sum of effective sample size was calculated as Neff = 103 136, with the sample prevalence set to 0.50 and the population prevalence to 0.05 [21].

### Lead SNP identification

We identified lead SNPs in each GWAS through clumping and thresholding using the same parameters used in the Big Five personality GWAS [14] with PLINK version 1.90b [34]. Specifically, we define a lead SNP as a common genetic variant (MAF ≥ 0.01) that is significant at *p* < 5×10^-8^ and independent of other lead SNPs (LD r² < .1) within a 250kb window. We aligned these SNPs to the 1000 Genomes Project Phase 3 European superpopulation [33] for the MR and colocalization analyses.

### Mendelian randomization

We performed two-sample MR analyses on the ADHD and Big Five lead SNPs to investigate bidirectional relationships between the ADHD GWAS and each personality trait GWAS [24], using the TwoSampleMR R package version 0.7.9 [35]. SNPs had to be present in both the ADHD and the personality GWAS.

Primary MR analyses were conducted using the Inverse Variance Weighted (IVW) method. We applied a Bonferroni correction for multiple testing (*p* < 0.05/(5x2), with 10 reflecting the number of bidirectional associations). The IVW method provides an overall estimate by regressing SNP–outcome associations on SNP–exposure associations, weighted by the inverse of their variance.

To assess the robustness of our findings and potential violations of MR assumptions, we performed several sensitivity analyses. First, we tested for heterogeneity across SNP-specific estimates using Cochran’s *Q* statistic. A significant heterogeneity statistic suggests that the effects of SNPs in the MR model vary more than expected, possibly due to confounding pleiotropy from third variables. However, heterogeneity alone does not necessarily imply confounding, as it may also reflect genuine differences in SNP effect sizes. To explore whether heterogeneity from the Cochran’s *Q* statistic was indeed driven by pleiotropy, we estimated the MR-Egger intercept. A significant non-zero intercept indicates the presence of directional (unbalanced) pleiotropy, which suggests that some SNPs may be associated with the outcome through third-variable pathways other than the intended personality-ADHD pathway.

Second, we performed an MR-Egger regression, which allows some SNPs in the MR analyses to not satisfy the strict MR assumptions. This method can provide unbiased estimates if the MR assumptions are violated by certain forms of directional pleiotropy, although it has reduced statistical power compared to the IVW method.

Third, we adopted a weighted median and mode approach. The median approach yields valid estimates of directional effects if at least half of the total weight of effects comes from valid SNP instruments. The weighted mode approach assumes that the largest cluster of SNP-specific effects represents the true effect.

Fourth, to assess the stability and reliability of our results, we examined whether individual lead SNPs exert a disproportionate influence on the MR results by performing leave-one-out analyses. To examine whether our estimates were affected by any single SNP, we iteratively repeated the MR analysis using the IVW method, omitting one lead SNP per iteration from the set of genetic instruments.

Fifth, we performed Mendelian randomization analyses using the MR-CAUSE (Causal Analysis Using Summary Effect Estimates) method (version 1.2.0.0335) [36]. Default package settings were used. Unlike the methods described above, which account for uncorrelated horizontal pleiotropy, MR-CAUSE leverages a broader genome-wide set of approximately independent variants while accounting for correlated pleiotropy arising from a heritable factor that influences both traits. It jointly estimates the causal effect and shared factor, then compares a sharing model with no causal effect against a causal model using the expected log pointwise predictive density (ELPD). Results are reported at nominal (*p* < 0.05) and Bonferroni (*p* < 0.05/10; absolute Z < 2.58) thresholds.

Finally, because both the deCODE and Yale-Penn-3 cohorts contributed to the personality and ADHD GWAS, we also conducted MR analyses with the MRlap R package version (version 0.0.3.3) [37] to test and correct for biases related to winner’s curse, weak instruments, and sample overlap in the IVW framework [38]. As done in the lead SNP identification step, pruning distance was set to 250 kb, pruning LD threshold to 0.1, and *p* < 5×10^-8^.

### Colocalization analyses

To obtain independent, variant-level support that MR associations were not solely driven by confounding LD, we conducted colocalization analyses between ADHD and the five personality trait GWAS using the coloc-SuSiE approach [30, 39], which accounts for multiple causal variants within a region by incorporating LD information.

Fine-mapping was performed separately in each GWAS on genomic loci defined by LD-based clumping of genome-wide significant SNPs (p < 5×10⁻⁸, r² < .10 within a 250 kb window, MAF ≥ 0.01) using PLINK version 1.90b [34], with overlapping clumped regions merged into single loci (excluding the Major Histocompatibility Complex [MHC]).

Each locus was fine-mapped using the *susie_rss* function from the susieR R package version 0.14.2 [30], supplying per-SNP Z scores, an LD matrix, and the median per-SNP sample size (N or effective N) for the locus. LD matrices were computed with PLINK version 1.90b [34] from the 1000 Genomes Project Phase 3 European superpopulation [33], restricted to SNPs with MAF ≥ 0.01, with the MHC region removed. Before fine-mapping, alleles were aligned to the reference panel, each locus was restricted to SNPs present in the reference panel, and loci with fewer than two SNPs were excluded. SuSiE objects were annotated with rsIDs and the locus LD matrix using the *annotate_susie* function for the colocalization analyses [39].

Credible SNP sets (each set with a 95% probability of containing the causal variant) identified by SuSiE were used to identify fine-mapped loci that overlapped in genomic coordinates between the ADHD GWAS and each personality trait GWAS. We then tested for colocalization between every pair of credible sets across the two traits using default prior probabilities (p1 = 1×10⁻⁴, p2 = 1×10⁻⁴, p12 = 5×10⁻⁶) with the *coloc.susie* function from the coloc R package version 5.2.3 [39]. Following *coloc.susie*’s default behavior, signal pairs were only colocalized if the variants shared between the two loci captured at least half of the posterior support for each trait’s credible set. Pairs below this threshold were automatically discarded.

A signal pair was said to colocalize when the posterior probability that both traits share a causal variant (H4) was equal to or greater than 0.80. We additionally report the conditional probability of a shared causal variant [H4/(H3+H4)], which quantifies the probability that the two traits share the same causal variant (H4) rather than having distinct causal variants in the region (H3), conditional on both traits being associated. Finally, we used the GWAS Catalog [40] to identify genes associated with the colocalizing SNPs and report their previous phenotype associations.

## RESULTS

### Genetic correlation analyses

The ADHD GWAS was positively genetically correlated with Neuroticism (*r*g = 0.32, SE = 0.03), Extraversion (*r*g = 0.18, SE = 0.03), and Openness (*r*g = 0.09, SE = 0.03), and was negatively correlated with Agreeableness (*r*g = -0.31, SE = 0.04) and Conscientiousness (*r*g = - 0.22, SE = 0.03; **Supplementary Table 1)**.

### Mendelian randomization

We conducted a two-sample MR analysis on all 36 lead SNPs for ADHD, and 703 out of 739 for Neuroticism, 158 out of 165 for Extraversion, 98 out of 104 for Openness, 88 out of 94 for Conscientiousness, and 27 out of 33 for Agreeableness (**Supplementary Tables 2-8; Supplementary Figure 1**).

When examining bidirectional effects between ADHD and Agreeableness (**Supplementary Tables 8-9; Figure 1**), we found a negative effect of ADHD liability on Agreeableness (IVW β = −0.05, SE = 0.01, *p* = 3.0×10⁻⁴). The weighted median estimate was also significant (β = −0.04, SE = 0.01, *p* = 0.003), while the weighted mode (β = −0.02, SE = 0.02) and MR-Egger (β = −0.05, SE = 0.07) estimates were directionally consistent but non-significant. The MR-Egger intercept was non-significant, indicating no evidence of directional pleiotropy. MR-CAUSE further supported a causal over a sharing model (ΔELPD = −4.68, z = −2.69).

**Figure 1.**
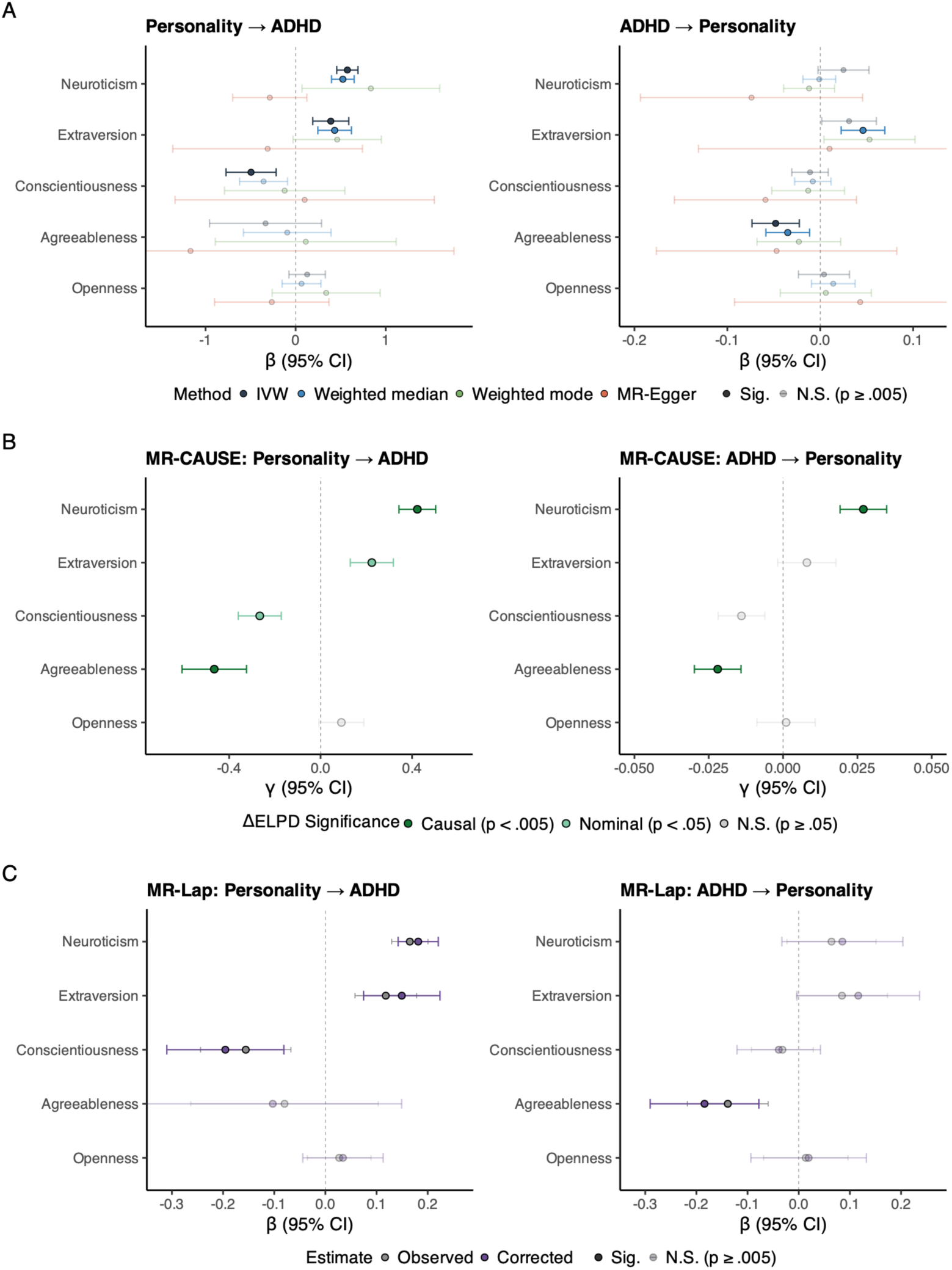
Bidirectional Mendelian randomization between ADHD and the Big Five personality traits. Effect estimates and their 95% confidence intervals (CIs). Multiple comparison Bonferroni correction for 10 tests (*p* < 0.05/10). Grey dots are Not Significant (N.S.). (A) Two-sample MR estimates (β) from four methods: inverse-variance weighted (IVW), weighted median, weighted mode, and MR-Egger. (B) MR-CAUSE estimates (γ); the causal model was favored over a shared/correlated-pleiotropy model in dark green at *p* < 0.005 and in light green at *p* < 0.05; Significance is based on the Z statistics of the difference in expected log pointwise predictive density (ΔELPD) between the two models. (C) MR-Lap estimates (β) before (observed) and after correction for sample overlap between the ADHD and personality GWAS (corrected).

MR-CAUSE also favored a causal effect of Agreeableness on ADHD (γ = −0.465, z = −3.08) that the instrument-based methods did not detect. This discrepancy may be due to the limited power of the Agreeableness instrument, which only included 27 variants and produced an imprecise estimate (IVW β = −0.33, SE = 0.32). Although MR-CAUSE results suggest that lower Agreeableness may increase ADHD liability, instrument-based analyses using larger GWAS are needed to test for causality.

When examining associations between Neuroticism and ADHD (**Supplementary Tables 8-9; Figure 1)**, we observed a robust effect of Neuroticism on ADHD liability. The IVW MR estimate indicated a positive effect (β = 0.57, SE = 0.06, *p* = 2.1×10⁻²¹), which was corroborated by the weighted median method (β = 0.52, SE = 0.06, *p* = 2.3×10⁻^16^). The weighted mode estimate was directionally consistent (β = 0.84, SE = 0.40) but did not survive multiple comparison correction (*p* = 0.032). We found evidence of heterogeneity across genetic instruments (Cochran’s *Q p* < 5.5×10⁻^124^), and a significant MR-Egger intercept (*p* = 2.2 × 10^-5^), indicating directional horizontal pleiotropy. The MR-Egger slope was imprecise and non-significant and the causal model was favored over the sharing model (ΔELPD = −5.42, z = −4.13) (**Supplementary Table 9**).

The causal model was also favored in the opposite direction, for ADHD on Neuroticism (ΔELPD = −5.86, z = −3.30; **Supplementary Table 9)**, although the directional association was not detected by instrument-based methods. The IVW estimate was directionally concordant but non-significant (β = 0.025, SE = 0.014, *p* = 0.08) and the weighted median, weighted mode, and MR-Egger estimates null. We found heterogeneity across instruments (Cochran’s Q *p* < 6.1×10⁻⁶¹), without directional pleiotropy. Given that the 36 ADHD instruments were appropriately powered to detect a similarly small but significant effect of ADHD on Agreeableness (β = −0.05, *p* = 3×10⁻⁴), the effect of ADHD on Neuroticism suggested by MR-CAUSE is likely to be small. We therefore interpret the MR-CAUSE signal as suggestive evidence of a potential small effect that requires further directional investigation.

Extraversion influenced ADHD liability (**Supplementary Tables 8-9; Figure 1**), with IVW estimates indicating a positive effect for Extraversion (β = 0.39, SE = 0.10, *p* = 1.3×10⁻⁴), consistent with the weighted median estimate (β = 0.43, SE = 0.10, *p* = 4.9×10^-6^). Although we found heterogeneity among the instruments (Cochran’s *Q p <* 2.0×10⁻^30^), the MR-Egger intercept was not significant, indicating that this heterogeneity is unlikely to reflect directional pleiotropy and more probably arises from balanced pleiotropy or genuine variation in SNP effects, consistent with the multidimensional nature of personality. MR-CAUSE also provided nominal support for a causal over a sharing model for Extraversion (ΔELPD = −4.23, z = −2.32), consistent in direction with the MR estimates. In contrast, non-significant results across the primary IVW and most sensitivity analyses indicated that ADHD was unlikely to have an effect on Extraversion (**Supplementary Tables 8-9; Figure 1)**. There was evidence of heterogeneity (Cochran’s *Q p <* 5.5×10⁻^23^) without directional pleiotropy (MR-Egger intercept), consistent with valid instruments. Although there was no evidence for a causal effect of ADHD on Conscientiousness, Conscientiousness was found to influence ADHD liability (**Supplementary Tables 8-9; Figure 1)**. IVW estimates indicated a negative effect for Conscientiousness (β = −0.50, SE = 0.14, *p* = 5.1×10⁻⁴), consistent with the weighted median estimate (β = −0.36, SE = 0.13, *p* = 8.8×10⁻³), which was near Bonferroni significance. Instrument heterogeneity was present (Cochran’s *Q p <* 1.4×10⁻^19^) but the MR-Egger intercept test was not significant. MR-CAUSE provided nominal support for a causal over a sharing model for Conscientiousness (ΔELPD = −3.94, z = −2.39).

Despite a small positive genetic correlation between Openness and ADHD, no tests indicated a significant influence of Openness on ADHD liability or vice versa.

Although there were some differences in the lead SNPs identified when using MR-Lap compared to PLINK (lead SNP overlap ranged from 78% to 94% across traits; **Supplementary Tables 2-7**), MR-Lap MR results were consistent with the primary MR results (**Supplementary Table 10; Figure 1**). We replicated the negative effect of ADHD liability on Agreeableness and Conscientiousness on ADHD, as well as the positive effect of Neuroticism and Extraversion on ADHD. The observed and corrected values of these associations differed significantly (*p* < 0.05/10), except for the association with Conscientiousness and ADHD. The MR-Lap-corrected effects were larger than the observed effect, suggesting that the uncorrected estimates were biased towards the null, likely due to weak instrument bias and regression to the mean [41]. In line with the primary results, the MR-Egger intercept test was significant for the effect of Neuroticism on ADHD (*p* = 2.9×10^-6^).

Leave-one-out analyses confirmed that no single SNP drove the results, suggesting that the main IVW results are robust to the included SNPs (**Supplementary Table 11**; **Supplementary Figure 2**).

### Colocalization analyses

SuSiE identified 2,739 credible sets across the six GWAS (ADHD, 22 credible sets at 20 loci; Agreeableness, 87 at 18; Conscientiousness, 271 at 70; Extraversion, 428 at 113; Openness,

292 at 69; Neuroticism, 1,639 at 372; **Supplementary Tables 12–17**). For ADHD, all 22 credible sets contained multiple variants (median set size = 19 SNPs, ranging from 2 to 149 SNPs). In contrast, most personality-trait credible sets resolved to a single variant (Agreeableness 77% [67/87]; Conscientiousness 66% [179/271]; Extraversion 73% [313/428]; Openness 71% [207/292]; Neuroticism 79% [1,289/1,639]; median set size = 1 SNP for every trait).

We tested 16 loci that overlapped in genomic coordinates between ADHD and the personality GWAS. Four were excluded from the colocalization analyses because too few credible-set SNPs were shared between datasets (all three Extraversion loci and one Neuroticism locus). Among the remaining loci, we found evidence of colocalization (H4 > 0.8) at three ADHD risk loci, comprising six colocalizing credible-set pairs across Agreeableness, Conscientiousness, and Neuroticism (**Supplementary Table 18**). The ADHD signal (rs9969232) colocalized with Agreeableness (rs1476535; H4 = 0.87) on chromosome 7. On chromosome 5, the ADHD signal (rs77960) colocalized with the same variant (rs2431108) in both Neuroticism (H4 = 0.99) and Conscientiousness (H4 = 0.99). A third region on chromosome 11 colocalized between ADHD and Neuroticism. However, only a single, poorly-resolved ADHD signal (rs10835362; PIP = 0.02, from a credible set with 104 variants) returned comparable support (H4 = 0.80–0.82) against three distinct Neuroticism variants (rs3935074, rs12576604, rs10835381), indicating an unresolved ADHD signal compatible with several Neuroticism signals rather than one well-defined shared variant.

Colocalization was generally sparse, likely reflecting the limited fine-mapping resolution of the ADHD GWAS. However, low ADHD PIP values should not necessarily be interpreted as evidence of weak colocalization (**Supplementary Table 18**). At the chromosome 5 and 7 loci, the conditional probability of a shared rather than distinct causal variant, H4/(H3+H4), remained high (≥ 0.87) despite low ADHD PIP values, and was reduced only at the weaker chromosome 11 locus.

Based on the GWAS Catalog, colocalized variants mapped to genes involved in psychiatric traits, cognitive function, and general health. The ADHD SNP rs9969232 and the Agreeableness SNP rs1476535 both mapped to FOXP2, a gene associated with neurodevelopment, educational attainment, and sleep. The ADHD SNP rs77960 colocalized with rs2431108 from Neuroticism and Conscientiousness, mapping to RNU6-334P and NIHCOLE, which are associated with mood disorders, depression, and stress-related traits. ADHD SNP rs10835362 colocalized with several Neuroticism SNPs (rs10835381, rs12576604, rs3935074) and mapped to LINC02758 and METTL15, which are involved in alcohol consumption, metabolism, and blood pressure regulation.

## DISCUSSION

This study leveraged the largest and most up-to-date genomic data of personality and ADHD to address a long-standing question of interest to researchers, clinicians, and parents alike: Does ADHD shape a child’s personality, or do certain personality traits make them more susceptible to developing ADHD? Using MR, alongside multiple sensitivity analyses, we find evidence of directional relationships between ADHD and personality that vary by personality trait. While three personality traits (higher Neuroticism and Extraversion, alongside lower Conscientiousness) increased genetic liability for ADHD, ADHD liability primarily influenced one personality trait: lower Agreeableness. These findings suggest that personality and ADHD are genetically linked in trait-specific and directional ways, offering a foundation for future work on how personality might inform our understanding of ADHD risk at the population level.

This study is the first to report an effect of ADHD liability on Agreeableness. The effect was consistent in direction and significance across the IVW, weighted median, leave-one-out, and MR-Lap analyses, with no evidence of directional pleiotropy, and was further supported by MR-CAUSE, which explicitly models correlated pleiotropy. Colocalization analyses added variant-level support, localizing a fine-mapped variant shared between ADHD and Agreeableness within the FOXP2 locus, a gene previously implicated in both traits [22, 42]. However, a single shared variant is unlikely to fully capture the shared genetic basis between the traits, and may instead reflect limited power to detect additional colocalizing loci [29]. Although MR-CAUSE also favored an effect in the reverse direction, whereby lower Agreeableness increased ADHD liability, better power GWAS are needed to test this hypothesis with instrument-based methods. The present results provide initial genetic evidence for a scar-model effect of ADHD on Agreeableness, consistent with the premise that psychiatric conditions can leave a lasting scar on personality [15]. In this context, lower Agreeableness may emerge as a consequence of ADHD, potentially reflecting interpersonal difficulties driven by impulsivity or emotional dysregulation [17], though these results warrant replication in larger samples.

Personality traits also appeared to influence ADHD risk. Across multiple analyses, including IVW, weighted median, leave-one-out, and MR-Lap, higher Neuroticism, higher Extraversion, and lower Conscientiousness were associated with greater ADHD liability. MR-CAUSE provided the strongest support for Neuroticism, suggesting that this association was unlikely to be explained by correlated pleiotropy. Evidence for Extraversion and Conscientiousness was weaker but directionally consistent and nominally significant. MR-CAUSE also favored an effect of ADHD on Neuroticism. However, this finding was not corroborated by the well-powered instrument-based analyses and therefore provides, at most, suggestive evidence of a small reverse effect that warrants replication when larger GWAS become available. Overall, our results more strongly support the directional effect of Neuroticism on ADHD liability.

Colocalization analyses provided complementary variant-level support for the associations of ADHD with Neuroticism and Conscientiousness, but not Extraversion. For Extraversion, the overlapping loci could not be evaluated because too few fine-mapped variants were shared across datasets. This likely reflects limited fine-mapping resolution rather than evidence against a shared signal. Together, these findings generally support the MR results, although additional shared variants may emerge as GWAS power and fine-mapping resolution improve.

Previous studies using less-powered GWAS have also reported an effect of Neuroticism on ADHD [27] and an influence of Extraversion on ADHD [28]. However, the association of ADHD on Extraversion, as well as the bidirectional relationships between ADHD and other personality traits, were not investigated, highlighting the novelty of our study.

The effects of personality on ADHD support the vulnerability model [1], suggesting that certain personality traits may increase an individual’s susceptibility to ADHD rather than arising as consequences of the disorder. Neuroticism, characterized by a heightened tendency to experience psychological stress, might predispose individuals to attentional difficulties through mechanisms such as cognitive overload [43] or task avoidance [44]. High Neuroticism could, in turn, serve as an early marker for children at genetic risk, with emotional distress and dysregulation contributing to the emergence of ADHD symptoms. Higher Neuroticism also predisposes to all other psychiatric disorders, including mood and anxiety disorders [22]. This explanation may be particularly relevant for the subgroup of children with ADHD who exhibit elevated levels of neuroticism [45]. Similarly, individuals with reduced Conscientiousness may struggle with goal-directed behavior, self-monitoring, and impulse control, which could exacerbate attentional difficulties and hyperactivity [2]. Conversely, children with high levels of conscientiousness may be less likely to be diagnosed with ADHD because they are able to compensate for their difficulties via this personality trait [1]. Finally, higher levels of sociability, impulsive engagement, or sensation-seeking tendencies, which are components of Extraversion, may also contribute to ADHD susceptibility, illustrating how certain personality traits typically considered adaptive might, in some contexts, increase vulnerability to attentional and behavioral difficulties. Supporting this, Extraversion has a positive genetic correlation with externalizing disorders and behaviors [46, 47], and a negative correlation with internalizing disorders [14].

Openness to experience was weakly genetically correlated with ADHD, which is mostly consistent with the null associations observed in phenotypic studies [4, 13]. Near-zero correlations suggest that ADHD and Openness may be largely independent at both the genetic and phenotypic levels. This finding supports Cheverud’s conjecture [48], which proposes that phenotypic and genetic correlations between complex traits are generally similar, although this concordance does not hold for all traits [49, 50]. When discrepancies between phenotypic and genetic correlations arise, they may reflect environmental influences shaping how traits manifest at the phenotypic level [50].

This study is not without limitations: First, the ADHD GWAS was conducted in people of European genetic ancestry, which limits the generalizability of our findings beyond European-ancestry populations. These results should be replicated in individuals from other genetic ancestry groups once the data become available. Second, we were unable to examine subdimensions of Neuroticism or distinct ADHD symptom domains, which may have differential associations with personality traits [4, 17]. Third, although personality traits show stability across development, the personality GWASs were based on adult samples and may not fully capture traits earlier in development. Moreover, since the ADHD GWAS included cases diagnosed in childhood, results may only generalize to early-onset and persistent forms of ADHD rather than to adult-diagnosed or late-emerging presentations. Importantly, however, the marked overlap between child temperament and adult personality traits suggests that the Big Five GWAS we used are appropriate instruments to model childhood personality [17]. Fourth, although the GWAS were conducted in sex-pooled samples, there is evidence that females tend to score higher on Conscientiousness, Agreeableness, and Neuroticism [51]. In light of these sex differences, future research should examine whether the directional associations between ADHD and personality traits vary by sex once sex-stratified GWAS of the Big Five traits become available. Fifth, the resolution of our MR and colocalization analyses was constrained by the genetic variants overlapping across traits, limiting both the number of loci available to test and the precision with which shared variants could be localized. Larger GWAS may identify more overlapping loci and resolve credible sets more finely, likely revealing additional shared variants, and, potentially, an effect of Agreeableness on ADHD. Finally, MR assumes that genetic instruments affect the outcome only through the exposure. We found evidence of directional pleiotropy only for the Neuroticism-ADHD association and even there, MR-CAUSE, which models uncorrelated and correlated pleiotropy, continued to support a causal effect. Nonetheless, residual pleiotropy through unmeasured pathways cannot be fully excluded and may bias our estimates.

Using the largest available genomic datasets, we find that ADHD genetic liability has limited effects on personality, decreasing Agreeableness, consistent with the scar model. In contrast, higher Neuroticism and Extraversion and lower Conscientiousness appear to increase ADHD liability, supporting the vulnerability model. These findings provide early, population-level insight into personality-related vulnerability to ADHD and motivate future research on whether personality traits could serve as behavioral indicators of genetic risk. Whether personality-informed support, such as emotion-regulation strategies for high Neuroticism or organizational-skills training for low Conscientiousness, could improve ADHD-related symptoms remains an open question for future work.

## STATEMENTS

### Author Agreement

All authors have reviewed and approved the final manuscript. We confirm that this work is original, has not been published before, and is not under consideration elsewhere. All authors had full access to all the data in the study and accept responsibility to submit for publication.

## Data Availability

No data was produced in the present study. This study exclusively uses publicly available, de-identified summary statistics from large-scale genome-wide association studies (GWAS). Public GWAS of each Big Five trait, excluding 23andMe participants, are available at OSF (https://doi.org/10.17605/OSF.IO/HGNSM) and the ADHD GWAS on the PGC website under "adhd2022" (https://pgc.unc.edu/for-researchers/download-results/).

## Acknowledgments

BC is supported by two French government grants managed by the Agence Nationale de la Recherche under the France 2030 program (reference ANR-22-EXPR0013 and ANR-23-IAHU-0010).

## Data availability

This study exclusively uses publicly available, de-identified summary statistics from large-scale genome-wide association studies (GWAS). All analyses were conducted on aggregate-level data with no individual-level identifiers, and no new data collection or direct participant involvement occurred. Summary statistics for GWAS of each Big Five trait, excluding 23andMe participants, are available at OSF (https://doi.org/10.17605/OSF.IO/HGNSM) and for ADHD on the PGC website under the "adhd2022" publication (https://pgc.unc.edu/for-researchers/download-results/).

## Ethical Information

The president of the Research Ethics Committee of Université Paris Cité (CER U-Paris Cité) certified that the above research project does not require submission to a research ethics committee under French law (Loi Jardé). The study uses only de-identified, summary-level data from genome-wide association studies (GWAS). No data were collected or produced for this study, and the investigators had no access to individual-level or identifiable participant data.

## Author contributions

HP and CMW designed the study. HP and CMW performed the analyses and drafted the manuscript. All authors discussed the results and contributed to the writing of the final manuscript.

