## Supplementary Information for "Testing Bidirectional Genetic Links Between ADHD and the Big Five Personality Traits"

### 1. **Figure S1 – Mendelian randomization (MR) scatter plots of bidirectional associations between ADHD and the Big Five personality traits.**

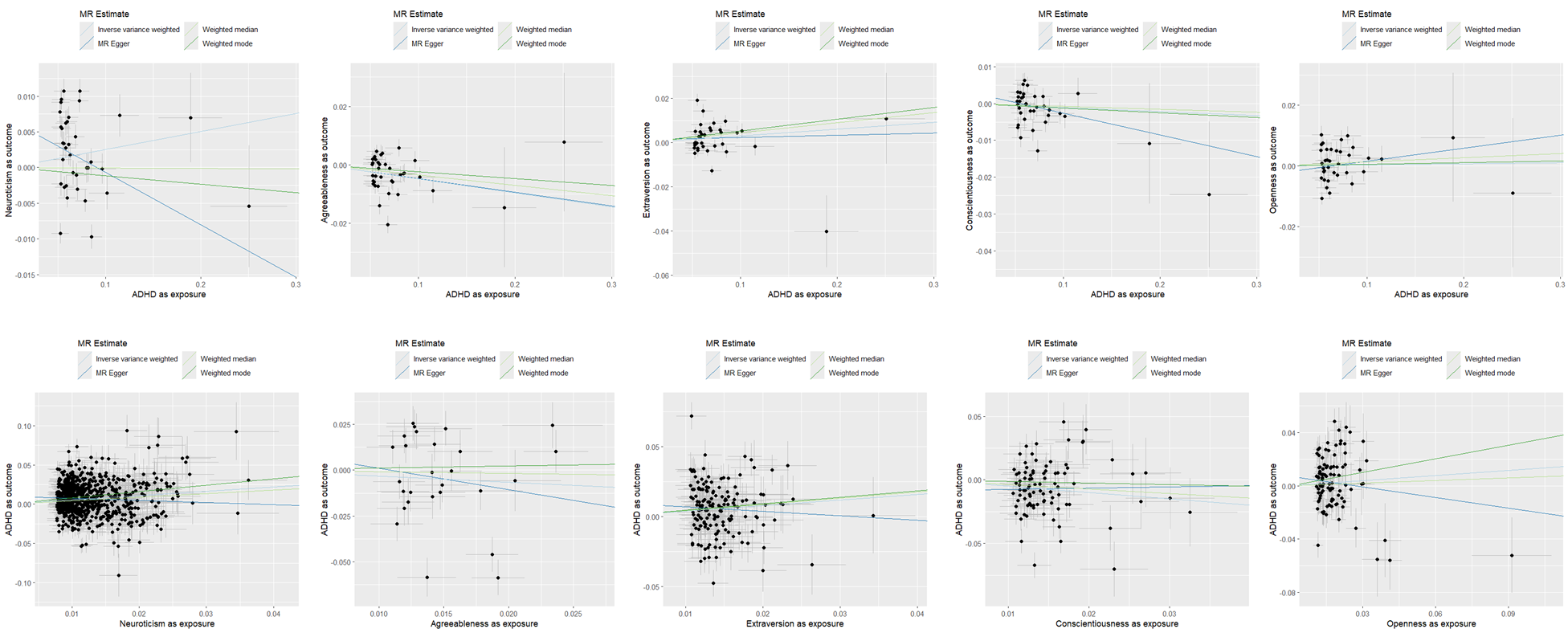

***Figure S1. Mendelian randomization (MR) scatter plots of bidirectional associations between ADHD and the Big Five personality traits.*** The top row shows ADHD liability as exposure and each personality trait as outcome (Neuroticism, Agreeableness, Extraversion, Conscientiousness, and Openness). The bottom row shows the reverse direction (personality traits as exposures and ADHD liability as outcome). Each point represents a lead SNP, along with its 95% confidence interval for the SNP-exposure association (x-axis) and the SNP-outcome association (y-axis). Fitted regression lines correspond to the IVW, MR-Egger, weighted median, and weighted mode estimators.

### 2. **Figure S2 – Leave-one-out Mendelian randomization (MR) analyses of bidirectional associations between ADHD liability and the Big Five personality traits.**

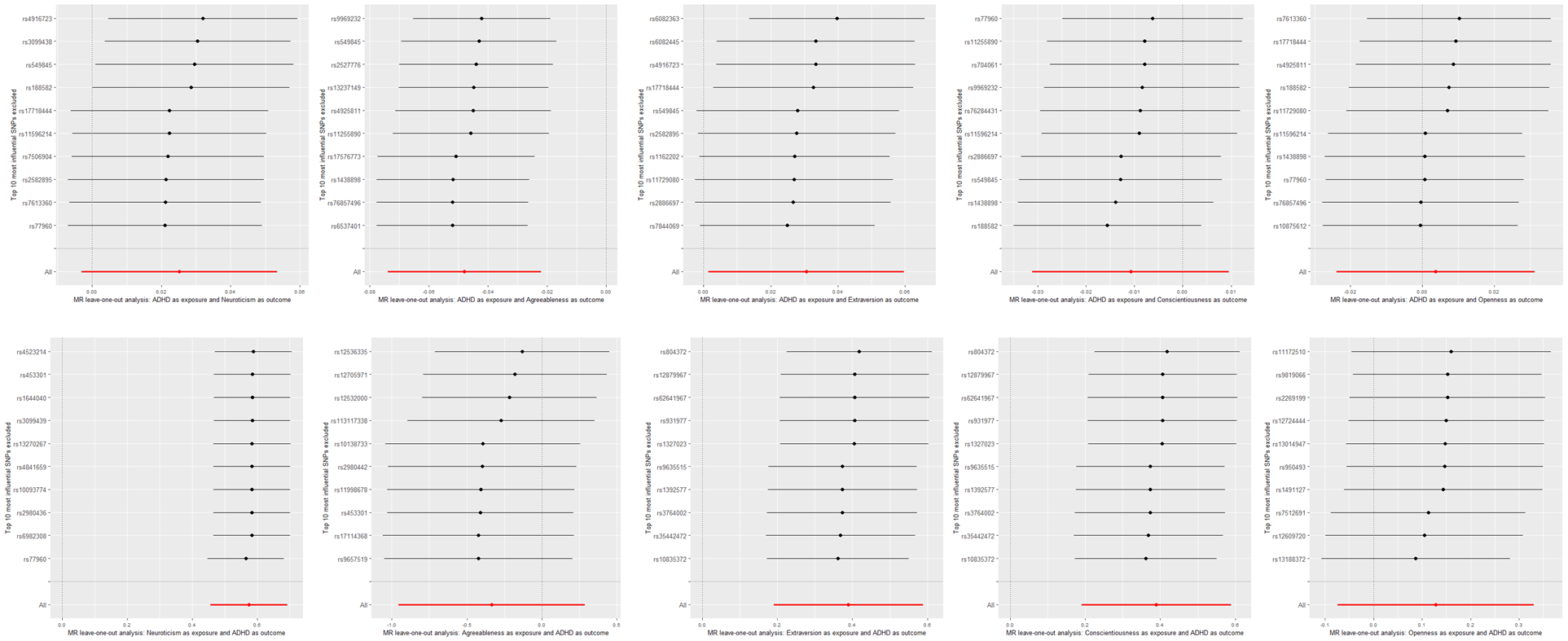

***Figure S2. Leave-one-out Mendelian randomization (MR) analyses of bidirectional associations between ADHD liability and the Big Five personality traits.*** The top row shows ADHD liability as exposure and each personality trait as outcome (Neuroticism, Agreeableness, Extraversion, Conscientiousness, and Openness). The bottom row shows the reverse direction (personality traits as exposures and ADHD liability as outcome). For each analysis, the MR estimate (using the IVW method) was iteratively recalculated while excluding one lead SNP at a time. The plots display the top 10 most influential SNPs excluded, illustrating whether any single variant disproportionately influenced the causal effect estimates.
